# AI for Mortality Prediction from Head Trauma Narratives

**DOI:** 10.1101/2025.02.20.25322619

**Authors:** Tuan D. Pham, Katya Marks, Declan Hughes, Domniki Chatzopoulou, Paul Coulthard, Simon Holmes

**Affiliations:** Barts and The London School of Medicine and Dentistry Queen Mary University of London, London, UK; Barts Health NHS Trust, London, UK; Department of Oral and Maxillofacial Surgery Royal London Hospital, London, UK

**Keywords:** Artificial intelligence, deep learning, natural language processing, head injuries, trauma, mortality, helicopter emergency medical services

## Abstract

Head injuries are a leading global cause of mortality and disability, highlighting the critical need for advanced prognostic tools to inform clinical decision-making and optimize healthcare resource utilization. For the first time, this study introduces a cutting-edge artificial intelligence (AI) framework designed to predict mortality outcomes from head injury narratives. Leveraging deep learning-based natural language processing techniques, the framework identifies and extracts key features from unstructured text describing injury mechanisms and patient conditions to train predictive models. Validation was conducted on a diverse dataset of 1,500 head injury cases using a stratified holdout approach, with 90% allocated for training and 10% for testing. The one-dimensional convolutional neural network model demonstrated strong performance, achieving averagely 85% accuracy, 74% correct mortality prediction, 88% correct survival prediction, and an impressive area under the receiver operating characteristic curve of 0.91. This work highlights the transformative potential of AI in harnessing narrative clinical data to enhance prognostic accuracy, paving the way for more effective, evidence-based management of head injury patients.

## 1 Introduction

Head injuries are a significant public health concern, contributing substantially to global mortality and long-term disability [1, 2]. According to the World Health Organization [3], neurological conditions, including traumatic brain injuries, are the leading cause of disability-adjusted life years and account for approximately 9 million deaths annually.

The complexity of head injuries, encompassing a wide spectrum of mechanisms, severities, and clinical presentations, poses significant challenges for timely and accurate prognostication [4, 5, 6, 7]. Effective prediction of outcomes, particularly mortality and neurological deterioration, is crucial for guiding clinical decisions, prioritizing interventions, and optimizing resource allocation in often resource-constrained healthcare environments.

In the emergency setting, rapid and precise prognostication of head injuries is critical for determining the urgency of surgical intervention, selecting appropriate neurocritical care strategies, and counseling patients if possible and their families regarding expected outcomes. Given the time-sensitive nature of emergency medicine, where every minute can influence survival and neurological recovery, accurate risk stratification is essential for prioritizing neurosurgical consultations and initiating timely interventions. Early identification of patients at high risk for deterioration enables clinicians to implement targeted management strategies, such as intracranial pressure monitoring [8], hyperosmolar therapy [9], or decompressive craniectomy [10], thereby improving overall patient outcomes.

Ultimately, improving prognostication in emergency medicine before surgery has profound implications for both individual patient outcomes and broader healthcare efficiency. By refining the ability to predict deterioration and surgical necessity, hospitals can allocate critical resources more effectively, reducing unnecessary interventions while ensuring that high-risk patients receive the urgent care they require. In an era of increasing healthcare demands, strengthening prognostic strategies is essential to advancing trauma care and improving survival rates for patients with severe head injuries.

The prognostication of head injuries in emergency medicine has garnered significant attention in recent years, particularly with the development of clinical decision rules and the evaluation of various risk factors that influence patient outcomes. One of the critical areas of focus has been the impact of anticoagulation therapy on head injury outcomes. Grewal et al. [11] conducted a population-based cohort study that highlighted the increased risk of intracranial hemorrhage among older patients on anticoagulants, particularly warfarin, following head injuries. Their findings suggested that these patients may experience worse outcomes compared to those not on anticoagulants, emphasizing the need for careful monitoring and management in emergency departments. Similarly, a systematic review by Minhas et al. [12] supported the recommendation for CT scanning in all anticoagulated patients with minor head injuries, regardless of symptoms, to mitigate the risk of missed intracranial injuries.

The evaluation of syncope as a precipitating factor for head injuries has also been explored. Furtan et al.[13] found that while syncope did not significantly increase short-term mortality, it was associated with higher long-term mortality rates. Their study corroborated the notion that older age, intracranial bleeding, and lower Glasgow Coma Scale (GCS) scores at admission are critical predictors of 30-day mortality in patients with head injuries. This aligned with findings from other studies that emphasized the importance of these factors in prognostication [14].

In pediatric populations, the use of clinical decision rules to guide imaging and management has been a focal point. The Pediatric Emergency Care Applied Research Network (PECARN) criteria, along with the Canadian Assessment of Tomography for Childhood Head Injury (CATCH) and the Children’s Head Injury Algorithm for the Prediction of Important Clinical Events (CHALICE), have been validated for their effectiveness in identifying children at risk for clinically significant traumatic brain injuries [15, 16, 17]. Bressan et al. [14] demonstrated that the PECARN criteria had the highest sensitivity in identifying significant head injuries, thus reducing unnecessary CT scans and associated radiation exposure. Furthermore, the study by Gizli et al. [15] compared these criteria and reinforced their utility in emergency settings, highlighting the need for selective imaging based on clinical presentation. The outcomes associated with different types of skull fractures in pediatric patients have also been investigated. Bressan et al. [18] found that isolated scalp hematomas in young children could be indicative of significant intracranial injuries, necessitating careful evaluation.

Emerging research has also focused on the prognostic value of physiological parameters in trauma patients. The reverse shock index multiplied by the GCS has been proposed as a potential predictor of mortality in severe trauma patients with head injuries, suggesting that integrating physiological metrics could enhance prognostic accuracy [19]. Furthermore, Ramdheen and Naicker [20] highlighted the Revised Trauma Score as a more accurate predictor of outcomes in patients with serious head injuries compared to traditional scoring systems.

Recent advances in artificial intelligence (AI) and natural language processing (NLP) have unlocked new possibilities for extracting and analyzing information from unstructured text [21, 22]. Deep learning-based NLP techniques, in particular, have demonstrated remarkable success in diverse healthcare applications, from automated diagnostics to predictive modeling [23, 24, 25, 26].

Traditional prognostic models for head injuries typically rely on structured data such as age, GCS scores, imaging findings, and other physiological parameters [27, 28, 29, 30, 31, 32]. While these approaches provide valuable insights, they often fail to capture the nuanced contextual information embedded in unstructured clinical narratives, such as the description of injury mechanisms, circumstances of the incident, and the initial presentation of the patient. These narrative data, routinely documented in medical records, hold untapped potential for improving prognostic accuracy.

This study represents a novel contribution as the first to develop an AI-powered framework specifically designed to analyze narrative descriptions of head injury cases for mortality prediction. Unlike previous approaches that primarily focus on structured datasets, our methodology integrates free-text clinical narratives, capturing subtleties that may not be fully reflected in numerical scoring systems. By incorporating real-world descriptions of injury mechanisms and initial clinical impressions, our framework expands the predictive capacity of AI models, making them more aligned with the complexities of head trauma management.

In this paper, we present the design, implementation, and evaluation of our AI framework, marking the first application of NLP-driven deep learning models for prognosticating head injury outcomes from unstructured clinical narratives. By leveraging the richness of narrative data, we demonstrate how this innovative approach can complement and enhance traditional prognostic models, offering a more holistic and accurate tool for head injury management. The findings of this study not only highlight the untapped potential of narrative-based AI models but also establish a new paradigm for integrating AI technologies into emergency and neurosurgical workflows. Our work pioneers an approach that could transform risk stratification, inform clinical decision-making, and ultimately improve patient outcomes in head trauma care.

## 2 Methods

### 2.1 Text data

A comprehensive dataset comprising narrative descriptions of head injuries from a cohort of 1,500 patients was obtained from London’s Air Ambulance Charity and utilized in this study. These narratives, recorded by pre-hospital emergency teams, provide detailed contextual information about injury mechanisms, initial clinical assessments, and early interventions. The dataset serves as a rich source of unstructured clinical text, offering insights beyond traditional structured data points.

Table 1 presents the demographic characteristics of the patient cohort along with the mechanisms of injury, illustrating the diverse causes and population distribution. Table 2 details the injury distribution and severity, highlighting the prevalence of various anatomical injuries and their clinical impact. These descriptive statistics provide a foundation for understanding the dataset composition, informing subsequent AI-driven prognostication efforts.

**Table 1:** Demographics and injury mechanisms. (SD = standard deviation, IQR = interquartile range, RTA = road traffic accident.)

|  |  |
| --- | --- |
| **Age** |  |
| Mean (SD) | 41.8 (19.9) |
| Median (IQR) | 38 (27, 56) |
| **Sex, $n$ (%)** |  |
| Female | 308 (20.5) |
| Male | 1192 (79.5) |
| **Mechanism, $n$ (%)** |  |
| Fall from ground level | 170 (11.3) |
| Fall down stairs | 186 (12.4) |
| Fall from height | 254 (16.9) |
| Assault with punches/kicks | 99 (6.6) |
| Assault with blunt weapon | 45 (3.0) |
| Chop injury | 3 (0.2) |
| Stab injury | 13 (0.9) |
| Gunshot wound | 6 (0.4) |
| RTA, car occupant | 90 (6.0) |
| RTA, motorcyclist | 84 (5.6) |
| RTA, cyclist or e-bike | 119 (7.9) |
| RTA, scooter or e-scooter | 7 (0.5) |
| RTA, pedestrian | 318 (21.2) |
| Fall from moving vehicle | 8 (0.5) |
| Hit by train | 29 (1.9) |
| Struck by falling or moving object | 21 (1.4) |
| Sporting injury | 2 (0.1) |
| Other | 3 (0.2) |
| Unknown | 43 (2.9) |
| **Penetrating injury, $n$ (%)** |  |
| No | 1473 (98.2) |
| Yes | 27 (1.8) |

**Table 2:** Injury distribution and severity.

|  |  |
| --- | --- |
| **Facial injury, $n$ (%)** |  |
| Orbit | 360 (24.0) |
| Zygoma | 289 (19.2) |
| Nose | 196 (13.1) |
| Maxilla | 312 (20.8) |
| Mandible | 65 (4.3) |
| **Head injury, $n$ (%)** |  |
| Basilar skull fracture | 1274 (84.9) |
| Vault skull fracture | 782 (52.1) |
| Extradural haematoma | 246 (16.4) |
| Subdural haematoma | 615 (41.0) |
| Subarachnoid haemorrhage | 716 (47.7) |
| Intracerebral haemorrhage | 105 (7.0) |
| Intraventricular haemorrhage | 93 (6.2) |
| Brainstem injury | 56 (3.7) |
| Cerebellar injury | 40 (2.7) |
| Diffuse axonal injury | 132 (8.8) |
| **Injury Severity Score** |  |
| Mean (SD) | 24.4 (12.4) |
| Median (IQR) | 22 (16, 30) |

Table 3 presents a selection of representative injury narratives, illustrating the detailed clinical descriptions recorded at the time of emergency care. Each example is accompanied by the corresponding patient outcome, categorized as either survival or mortality following surgical intervention. These narratives highlight the variability in injury mechanisms, severity, and initial clinical assessments, providing valuable context for understanding how unstructured text data can contribute to prognostic modeling.

**Table 3:** Narratives and outcome of head injuries.

| Patient # | Descriptions | Outcome |
| --- | --- | --- |
| 1 | base skull fx - complex; comminuted; ring; hinge; open w/ tissue lossface skin/subcutaneous/muscle abrasioncerebrum hematoma epidural/extradural - tinycerebrum pneumocephaluscerebrum hematoma intracerebral NFSvault skull fx - comminuted; compound w/ dura | alive |
| 2 | pneumothorax NFSlung contusion - bilateral NFSadrenal gland inj NFSliver inj NFSscalp lac; minor; superficialdiffuse axonal injury w/ LOC >24hrs NFScerebrum subarachnoid hemorrhage w/ coma <6hrscerebrum intraventricular hemorrhagecerebrum single contusion | dead |
| 3 | vault skull fx - closed; simple; undisplaced; diastatic; linearorbit fx - inferior orbital rim - closed or NFSbase skull fx w/o CSF leaknose fx - closed or NFSmaxilla fx | alive |
| 4 | cerebrum hematoma - subdural - large; massive; extensivecerebrum hematoma - intracerebral - largecerebellum subarachnoid hemorrhagebase skull fx w/o CSF leakvault skull fx - closed; simple; undisplaced; diastatic; linear | dead |
| 5 | base skull fx NFScerebrum subarachnoid hemorrhage NFScerebrum hematoma epidural/extradural NFScerebrum multiple contusion bilateral NFSvault skull fx NFS | alive |
| 6 | cerebrum multiple contusion bilateral - extensive; massivebase skull fx NFScerebrum subarachnoid hemorrhage w/ coma >6hrscerebrum hematoma - subdural - large; massive; extensive | dead |
| 7 | base skull fx w/ CSF leakvault skull fx NFSpneumothorax NFSscapula fx - bodycerebrum multiple contusion unilateral NFScerebrum hematoma - subdural - small; moderate - bilateralmetacarpus fx - lateral fingerdistal ulna fx - extra-articularradius shaft fx | alive |

This study was approved by the Research Ethics Committee of Queen Mary University of London (QME25.0913).

### 2.2 Deep learning of textual information

Building on a prior study on text-based classification of pediatric dental diseases from panoramic radiographs [26], this study explores three deep learning models for processing narratives of head injuries. These are long short-term memory (LSTM) network [33], bidirectional encoder representations from transformers (BERT) [34], and one-dimensional convolutional neural network (1-D CNN) [35].

LSTM networks are a specialized type of recurrent neural network (RNN) designed to handle sequential data more effectively by addressing the vanishing gradient problem, a common limitation in traditional RNNs. LSTMs excel at learning long-range dependencies, making them particularly well-suited for tasks involving physiological signal analysis [36, 37], NLP [38, 39], and speech recognition [40, 41]. Their ability to retain relevant information over extended sequences has made them a widely used architecture in applications requiring contextual understanding over time.

Unlike standard RNNs, which struggle to preserve information over long sequences, LSTMs incorporate a gating mechanism that regulates the flow of information through the network. The model consists of three key gates: the forget gate, which determines which past information should be discarded; the input gate, which decides what new information to store in the cell state; and the output gate, which selects the information to pass to the next time step. These gates work collectively to enable the network to selectively remember or forget information, effectively capturing long-term dependencies while mitigating gradient decay issues. This structure makes LSTMs particularly advantageous for modeling complex sequential patterns, such as clinical narratives in medical datasets.

BERT is a deep learning model designed to process and understand natural language with high contextual awareness. This model represent words independently of their context, and captures deep bidirectional contextual relationships by considering both preceding and following words in a sentence. This bidirectionality allows BERT to generate context-sensitive word representations, making it particularly effective for complex NLP tasks such as text classification [42], named entity recognition [43], question answering [44], and natural language inference [45, 46].

At its core, BERT is built on the transformer architecture, which utilizes self-attention mechanisms to process entire sequences in parallel rather than sequentially, as seen in RNNs and LSTM networks. This design enables BERT to capture long-range dependencies between words more efficiently, leading to superior performance in language understanding tasks. Unlike traditional sequence-based models, which process text in a left-to-right or right-to-left manner, BERT applies bidirectional encoding, ensuring a more comprehensive understanding of sentence structure and meaning.

BERT was pretrained on large-scale text corpora using two key self-supervised learning objectives: masked language modeling (MLM) and next sentence prediction (NSP). In MLM, a percentage of words within an input sentence are randomly masked, and the model learns to predict these missing words based on the surrounding context. This forces BERT to develop a deep contextual understanding of word relationships. In NSP, the model was trained to determine whether a given sentence logically follows another, helping it grasp discourse-level relationships between sentences. These pretraining strategies enable BERT to generalize effectively across diverse NLP tasks, requiring only minimal fine-tuning on domain-specific datasets to achieve state-of-the-art performance.

By leveraging its rich contextual representations and scalable transformer-based architecture, BERT has significantly advanced NLP research and applications, outperforming previous models in various benchmark evaluations. Its adaptability to domain-specific fine-tuning makes it a powerful tool for specialized text analysis, including clinical and biomedical applications where nuanced language understanding is critical.

For text classification using a 1D-CNN in this study, the network architecture is specifically designed to handle textual input and capture local dependencies within sequences. The model begins with an input layer, corresponding to the channel dimension of the input integer sequence. This ensures compatibility with the convolutional layers, which operate over sequential data. The input data are then embedded using a word embedding layer with a dimension of 50, mapping discrete tokens into continuous vector representations that preserve semantic relationships between words.

To extract contextual patterns from the text, multiple convolutional blocks are constructed, each designed to capture different *n*-gram features representing varying contextual dependencies in the data. Specifically, four convolutional blocks are created, corresponding to *n*-gram lengths of 2, 3, 4, and 5. Each block follows a standardized structure, consisting of the following components.

(a) A 1D convolutional layer, which applies 100 filters of size corresponding to the *n*-gram length, enabling the network to detect key local features from the input text.
(b) Batch normalization, which stabilizes the learning process by normalizing activations, improving convergence speed, and reducing internal covariate shift.
(c) A ReLU activation layer, which introduces non-linearity, allowing the model to learn complex patterns in textual data.
(d) A dropout layer with a rate of 0.2, serving as a regularization technique to prevent overfitting by randomly deactivating a fraction of neurons during training.
(e) A global max pooling layer, which reduces dimensionality by extracting the most salient feature from each filter, ensuring that the most informative patterns are retained.

Each of these convolutional blocks operates independently on the word embedding layer, extracting hierarchical representations of the input text. The outputs from all blocks are then concatenated using a concatenation layer, combining the learned features from different *n*-gram contexts into a unified representation. This architecture allows the model to capture both short-range and long-range dependencies within the input sequences, enhancing its ability to differentiate between textual patterns relevant to classification.

In the final stage, the concatenated feature representation is passed through a fully connected layer, which maps the extracted features to class predictions. A softmax layer is then applied to produce probability distributions over the target classes, enabling the network to make categorical predictions. The overall architecture is structured to efficiently handle varying *n*-gram lengths, ensuring that the network remains robust to different textual structures.

## 3 Results

The LSTM model architecture used in this study had a sequence input layer configured with an input size of 1, followed by a word embedding layer with an embedding dimension of 50, which transformed input tokens into dense vector representations. The LSTM layer with 100 hidden units was included, set to output only the final hidden state (last mode) to process the entire sequence. To facilitate classification, a fully connected layer was added, where the number of units corresponded to the number of classes in the dataset. To enhance generalization and prevent overfitting, a dropout layer with a rate of 0.2 was applied. Finally, a softmax layer was used to classify the input into one of the two target classes (survival or mortality).

For training, the Adam optimizer was selected due to its computational efficiency and adaptive learning rate properties, which helped accelerate convergence while maintaining stability. The cross-entropy loss function was used as the objective function, as it was well-suited for classification tasks by measuring the difference between predicted and actual probability distributions. The training process was conducted over a maximum of 600 epochs, allowing the model sufficient time to learn complex patterns in the data.

A batch size of 128 was specified to balance computational efficiency and generalization, ensuring stable gradient updates while maintaining a reasonable memory. To assess model performance, accuracy was chosen as the primary evaluation metric, providing a straightforward measure of classification correctness. Additionally, validation data was incorporated into the training pipeline, enabling real-time monitoring of generalization performance. The model’s weights were saved at the epoch yielding the best validation performance, preventing overfitting and ensuring that the final model retained optimal predictive capability.

The BERT-Base mode, comprising 108.8 million trainable parameters, was utilized for the prediction task in this study. To process the textual data, a tokenizer was employed to convert the raw text into sequences of integers, allowing the model to interpret and encode the input. The tokenization process ensured that the text was formatted in a manner compatible with BERT’s architecture, including padding shorter sequences and truncating longer ones to maintain consistency across input samples.

After tokenization, the dataset was partitioned into training and validation sets. To optimize training efficiency, the tokenized sequences were organized into mini-batches, which facilitated parallel computation and efficient gradient updates. Mini-batching is a crucial step in training large-scale models like BERT, as it balances computational efficiency with memory constraints. Once the text data was prepared, the BERT model transformed the tokenized input into feature vectors, extracting rich contextual embeddings that encapsulated the semantic meaning of the text. These embeddings served as input features for both the training and validation datasets, forming the foundation for the downstream prediction task.

To classify the BERT-derived feature vectors, a deep learning classification network was constructed. The architecture included a feature input layer, followed by a fully connected layer responsible for mapping the extracted feature vectors to class labels. A dropout layer was incorporated to reduce overfitting by randomly deactivating a fraction of the neurons during training, ensuring better generalization to unseen data. Finally, a softmax layer was used to generate probability distributions over the target classes, enabling the model to produce interpretable classification outputs. This classification network was specifically designed to leverage the deep contextual representations learned by BERT, enhancing the model’s ability to distinguish between outcome classes.

BERT’s training process was configured to ensure optimal model performance and stability. A mini-batch size of 128 was chosen to balance computational efficiency with convergence speed. The Adam optimizer was employed to adjust model parameters dynamically, leveraging adaptive learning rates for faster and more stable optimization. Training was conducted over a maximum of 600 epochs, allowing the model sufficient time to learn from the data while avoiding premature convergence. An initial learning rate of 0.0001 was selected to enable gradual weight updates, reducing the risk of overshooting local minima. To maintain a consistent data order and avoid introducing variability due to random permutations, data shuffling was disabled during training. This configuration ensured that the model was exposed to data in a controlled manner, facilitating stable convergence and reproducible results.

For the 1D-CNN model, training was conducted using the Adam optimizer, which was chosen for its adaptive learning rate properties and efficient gradient-based optimization, ensuring fast and stable convergence. A mini-batch size of 128 was used to balance computational efficiency and training stability, allowing for smoother gradient updates while maintaining a manageable memory footprint.

To optimize the model’s predictive performance, cross-entropy loss was employed as the objective function, as it is well-suited for classification tasks and provides a measure of how well the predicted probability distribution aligns with the actual class labels. The training process included a validation phase, where a separate validation dataset was used to monitor the model’s generalization ability and detect potential overfitting. After each training epoch, the model’s performance on the validation set was assessed, and the network with the lowest validation loss was saved to ensure that the final model retained optimal generalization capability rather than overfitting to the training data.

The dataset contains 17 instances with missing class labels, which were treated as “NaN” (not a number) to indicate the absence of a valid classification. To ensure the integrity of model training and evaluation, these missing data points were excluded from the analysis. Discarding incomplete records prevents potential biases or inaccuracies that could arise from imputed values, particularly in a high-stakes domain like mortality prediction. Additionally, preprocessing steps ensured that only complete and reliable data were used, optimizing the AI models’ ability to learn meaningful patterns from the available information.

Figure 1 presents histograms illustrating the class and word length distributions within the training dataset, providing insights into data imbalance and text complexity. Similarly, Figure 2 visualizes the most frequently occurring words in the training and test datasets through word clouds, highlighting key patterns and linguistic features within the text. Figure 3 shows the training and validation/testing processes of the three AI models.

**Figure 1:**
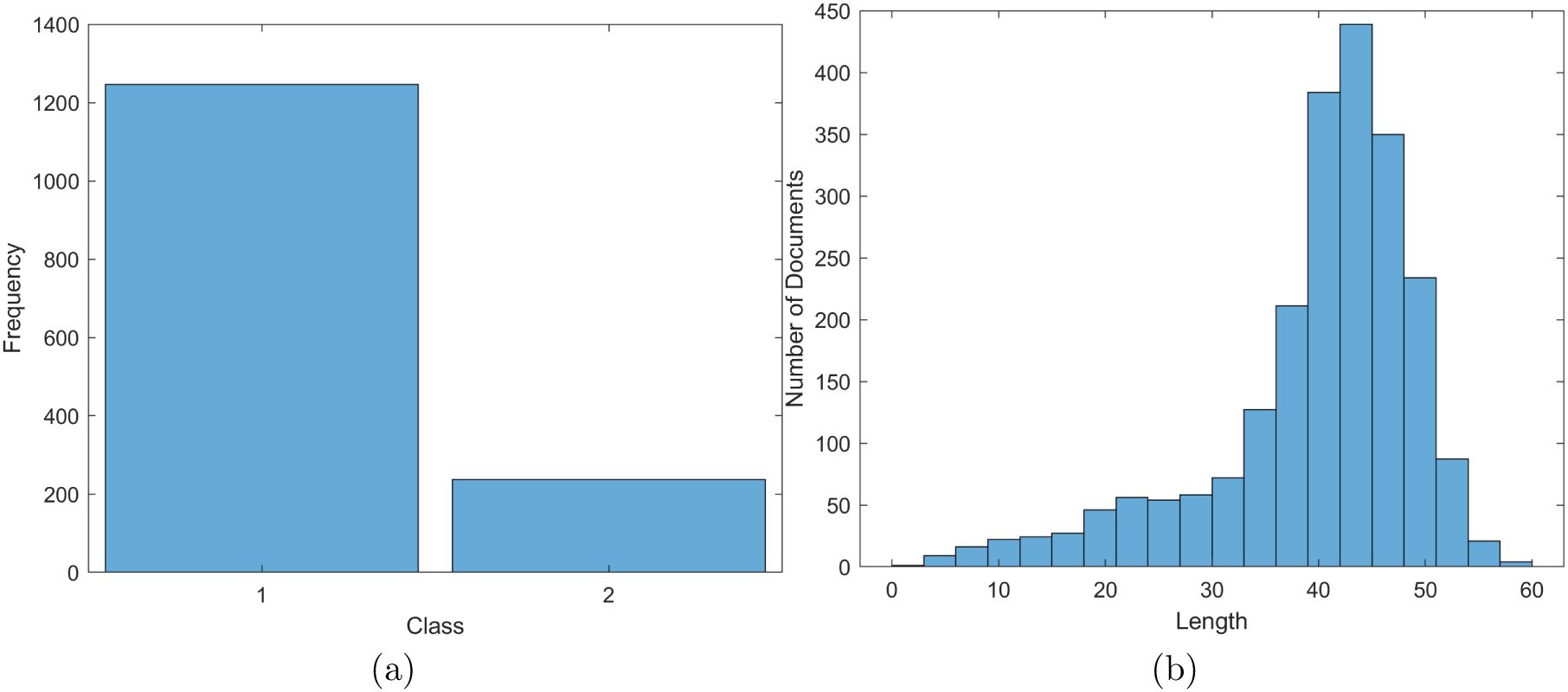
Histograms of class distribution (a), where classes 1 and 2 denote survival and mortality, respectively; and word lengths of training data (b).

**Figure 2:**
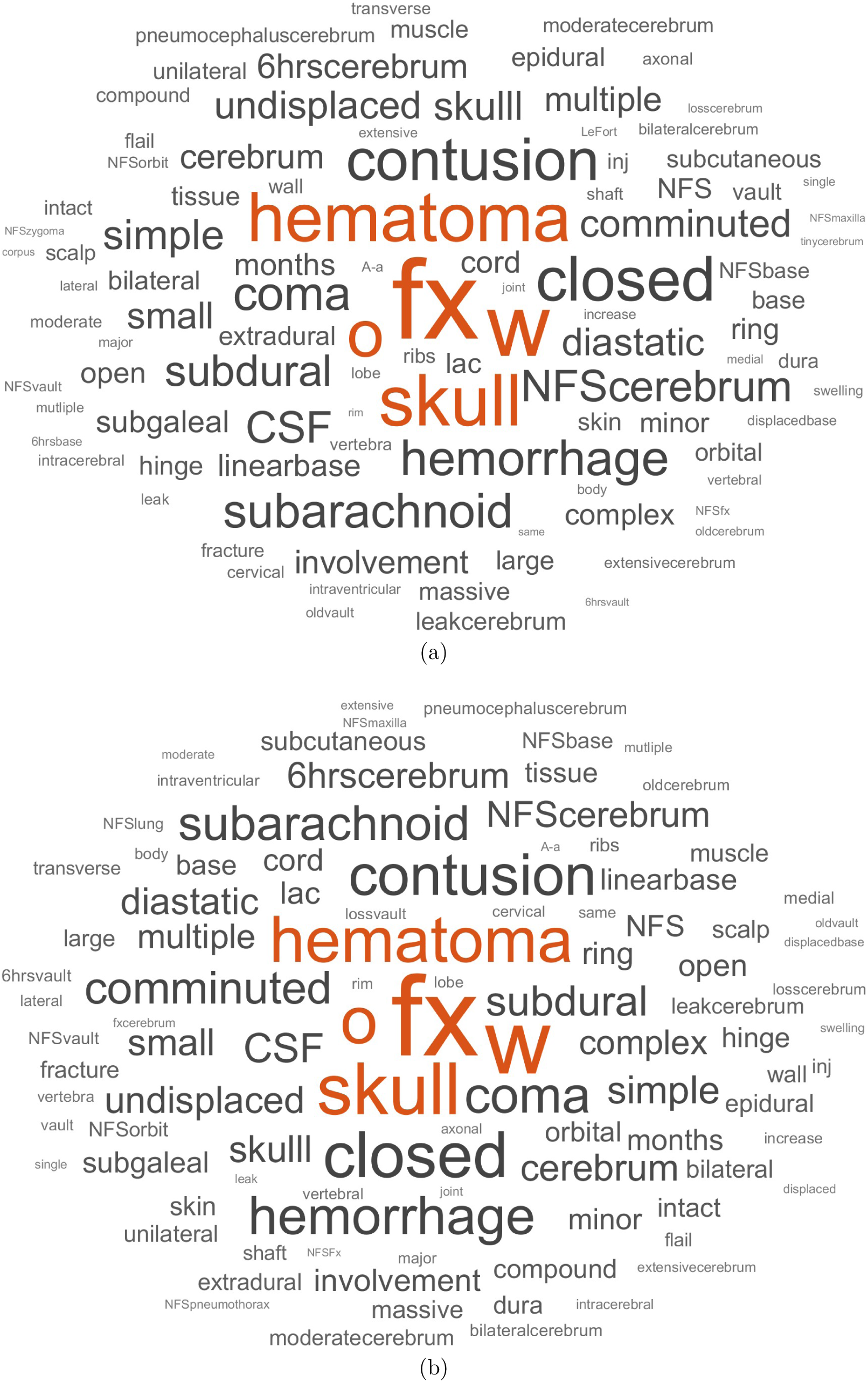
Word clouds of training (a) and test (b) data.

**Figure 3:**
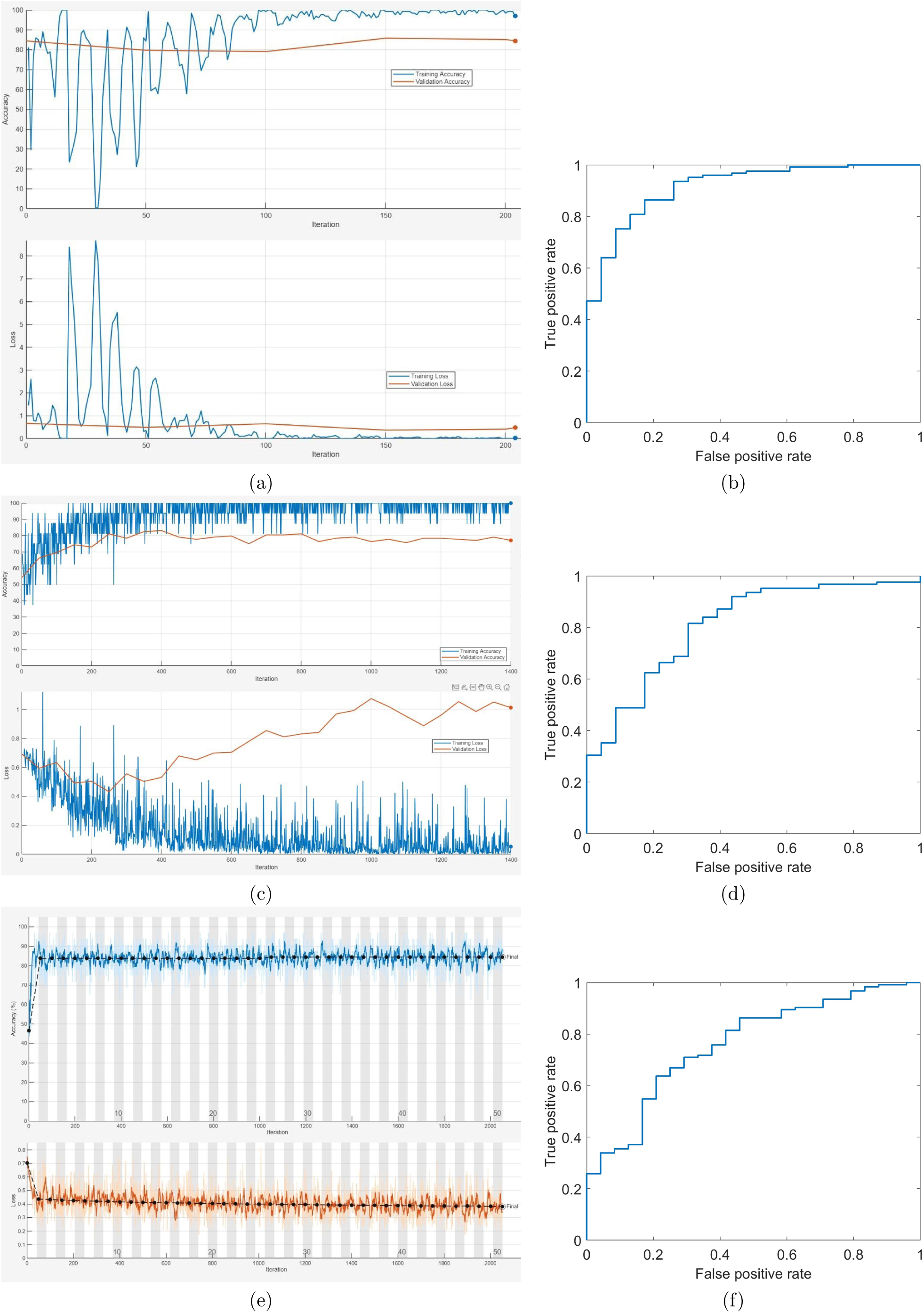
Training and validation processes and AUCs: 1D-CNN (a) and (b), LSTM (c) and (d), and BERT feature based net (e) and (f), where solid and dotted lines indicate training and validation, respectively.

Table 4 shows the performance of the three AI models (LSTM, BERT, and 1D-CNN), which was evaluated using key classification metrics, including accuracy (ACC), sensitivity (SEN), specificity (SPE), precision (PRE), F1-score (F1), and area under the curve (AUC). These metrics provide a comprehensive assessment of each model’s ability to predict mortality (SEN) and survival (SPE) while also measuring their overall discriminative power. These metrics, with the exception of AUC, are defined in Table 5 for reference. AUC is a measure of the model’s overall discriminative ability.

**Table 4:** Performance metrics of AI models.

| Model | ACC (%) | SEN (%) | SPE (%) | PRE (%) | F1 | AUC |
| --- | --- | --- | --- | --- | --- | --- |
| LSTM | $81.08 \pm 0.00$ | $60.87 \pm 0.00$ | $84.80 \pm 0.00$ | $42.42 \pm 0.00$ | $0.50 \pm 0.00$ | $0.81 \pm 0.00$ |
| BERT | $85.82 \pm 1.91$ | $17.30 \pm 18.57$ | $98.80 \pm 1.70$ | $85.00 \pm 21.21$ | $0.25 \pm 0.24$ | $0.79 \pm 0.03$ |
| 1D-CNN | $85.47 \pm 0.48$ | $73.91 \pm 0.00$ | $87.60 \pm 0.57$ | $52.32 \pm 1.14$ | $0.61 \pm 0.01$ | $0.91 \pm 0.00$ |

**Table 5:** Performance measures used for evaluating the binary classification AI models. (TP = true positive, mortality cases correctly identified as mortality; FP = false positive, survival cases incorrectly classified as mortality; TN = true negative, survival cases correctly identified as survival; FN = false negative, mortality cases incorrectly classified as survival.)

| Measure | Equation |
| --- | --- |
| Accuracy (ACC) | $\frac{TP+TN}{TP+TN+FP+FN}$ |
| Sensitivity (SEN) | $\frac{TP}{TP+FN}$ |
| Specificity (SPE) | $\frac{TN}{TN+FP}$ |
| Precision (PRE) | $\frac{TP}{TP+FP}$ |
| F1 score | $2 \times \frac{PRE \times SEN}{PRE+SEN}$ |

## 4 Discussions

In terms of sensitivity, which reflects the model’s ability to correctly identify mortality cases, the 1D-CNN model achieved the highest score at 73.91%, indicating that it was the most effective at detecting mortality. In contrast, BERT exhibited the lowest sensitivity at 17.30%, suggesting that it struggled to capture mortality-related patterns despite its strong performance in other areas. The LSTM model had a moderate sensitivity of 60.87% but was outperformed by 1D-CNN. Specificity, which is crucial for accurately predicting survival cases, showed a different trend. BERT demonstrated exceptional specificity at 98.80%, making it highly reliable in identifying patients who survived. Both LSTM and 1D-CNN also performed well in this regard, with specificity scores of 84.80% and 87.60%, respectively. However, the stark contrast between BERT’s high specificity and low sensitivity highlights a major limitation, while it is highly confident in predicting survival, it fails to identify a significant portion of mortality cases. In contrast, 1D-CNN achieved a better balance between sensitivity and specificity, making it a more reliable choice for real-world applications where both types of predictions are critical.

Precision, which measures how many of the predicted mortality cases were actually correct, further underscores the differences between the models. BERT recorded the highest precision at 85.00%, indicating that when it did predict mortality, it was highly confident in its predictions. However, this high precision came at the cost of extremely low sensitivity, meaning that it missed many true mortality cases. The LSTM model had the lowest precision at 42.42%, while 1D-CNN demonstrated a better balance with a precision of 52.32%. Similarly, the F1-score, which represents the harmonic mean of precision and sensitivity, was highest for 1D-CNN at 0.61, followed by LSTM at 0.50. BERT had the lowest F1-score at 0.25, confirming that its high precision was offset by poor recall of mortality cases. From a clinical perspective, the 1D-CNN model is the most suitable for real-world application, as it provides the best balance between mortality detection and survival classification.

The first application of AI for mortality and survival prediction using narrative descriptions of head injury cases addressed in this study represents a groundbreaking advancement in trauma care and emergency medicine. this AI-driven approach leverages unstructured text data from pre-hospital and emergency department records. These free-text narratives, often recorded by paramedics and trauma teams, contain rich contextual information about injury mechanisms, initial assessments, and clinical impressions that structured data alone may fail to capture. Key innovations and practical benefits include the following.

(a) *Enhanced early prognostication:* AI-driven NLP models can extract critical details from narrative descriptions that may indicate the severity of injury or likelihood of deterioration. By recognizing subtle linguistic patterns, such as descriptions of altered consciousness, hypotension, or the mechanism of injury, AI models can provide real-time risk stratification, helping clinicians anticipate high-risk cases before imaging or laboratory results become available.
(b) *Improved triage and resource allocation:* In emergency and pre-hospital settings, accurate risk assessment is crucial for prioritizing interventions and optimizing resource utilization. AI-driven mortality prediction from text can assist in identifying critically injured patients who require immediate neurosurgical evaluation, ICU admission, or transfer to a specialized trauma center. Conversely, it may help avoid unnecessary escalations of care for lower-risk patients, thus reducing overcrowding in emergency departments and ensuring efficient use of hospital resources.
(c) *Reducing subjectivity in clinical decision-making:* Trauma assessment often involves clinician-dependent judgments, which may vary based on experience, cognitive biases, or incomplete information at the time of evaluation. AI models trained on large datasets can provide objective, data-driven insights to support clinical decision-making, reducing variability and improving consistency in mortality and survival predictions.
(d) *Potential for integration with electronic health records:* As hospitals and trauma centers increasingly adopt electronic health record systems, AI-powered NLP models can be integrated to automatically analyze and summarize narrative reports, flagging high-risk cases in real time. This integration can improve clinical workflow efficiency, ensuring that critical cases receive prompt attention from trauma teams.
(e) *Facilitating retrospective analysis and quality improvement:* AI-driven analysis of historical trauma narratives enables researchers and healthcare administrators to identify patterns in head injury outcomes, assess variations in clinical practice, and develop evidence-based guidelines for head trauma management. By continuously refining AI models with new data, predictive accuracy and reliability can improve over time, leading to better patient outcomes.

Despite its potential, the AI-driven approach to predicting mortality and survival based on head injury narratives has several limitations, outlined below.

(a) *Data quality and variability:* Clinical narratives are often inconsistent, subjective, and influenced by differences in documentation styles among healthcare providers, which may affect model reliability. In emergency situations, the data entry clerk quickly reviews patient records and identifies important keywords in the transcribed notes. By recognizing key terms related to medical conditions or treatments, they help the system organize critical patient information. When the data are further refined by including details on severity, a deeper level of analysis becomes possible. This added detail would enhance the accuracy of AI models, allowing them to make more precise predictions and support better clinical decision-making in urgent care scenarios.
(b) *Limited generalizability:* The model was trained on data from a specific trauma system (London’s Air Ambulance), and its performance may vary across different regions, hospitals, or emergency settings with different documentation practices.
(c) Imbalance in mortality cases: Mortality cases are typically less frequent than survival cases, potentially leading to biased predictions that favor the majority class.
(d) *Lack of multimodal integration:* Current models rely solely on text data, missing out on complementary information from imaging, physiological parameters, and laboratory results that could enhance predictive accuracy.
(e) *Explainability and interpretability:* AI models, especially deep learning-based approaches, often function as “black boxes”, making it difficult to interpret their reasoning, which may limit clinical trust and adoption.

Given the limitations mentioned above, future research may focus on the following areas.

(a) *Improving data standardization:* Developing NLP techniques to handle inconsistencies in clinical narratives can enhance robustness.
(b) *Cross-center validation:* Testing the model across multiple trauma centers and healthcare systems can improve generalizability.
(c) *Multimodal AI models:* Combining textual data with imaging and physiological parameters can lead to more comprehensive and accurate prognostication.
(d) *Bias mitigation strategies:* Implementing techniques such as data augmentation and cost-sensitive learning can address class imbalance issues.
(e) *Enhancing model interpretability:* Using explainable AI approaches can make predictions more transparent and clinically interpretable, facilitating adoption in real-world settings.

## 5 Conclusion

This study represents a pioneering application of AI in mortality and survival prediction using narrative descriptions of head injury cases. By leveraging unstructured clinical text, the model captures nuanced details often overlooked in structured data, enhancing early prognostication and decision-making in emergency settings. The results demonstrate that deep learning-based NLP models, particularly 1D-CNN, can achieve a strong balance between sensitivity and specificity, making them suitable for real-world deployment in trauma care. The findings highlight the potential of text-based AI systems to support clinicians by providing objective risk assessments that can improve triage efficiency and optimize resource allocation.

Despite its promise, challenges such as variability in clinical documentation, data imbalance, and the need for broader validation remain. Future research should focus on integrating multimodal data sources, improving model interpretability, and ensuring adaptability across different healthcare settings. By addressing these challenges, AI-driven prognostic models can become valuable tools in trauma care, contributing to more timely interventions and better patient outcomes.

## Statements and Declarations

### Competing Interests

All authors declare no conflict of interest.

### Author contribution

TDP contributed to the conception of using text for AI-based prognostication, technical design, computer coding and implementation, and writing the original manuscript. SH, DC, and PC proposed the research question on mortality prediction using AI. KM and DH conducted the descriptive statistical analysis. All authors contributed to the review, data analysis and interpretation, and approved the final manuscript.

### Data availability

The fully anonymized dataset used in this study is available at the first author’s personal website: https://sites.google.com/view/tuan-d-pham/codes under the title “Prognostication of head injuries”.

### Software availability

MATLAB codes implemented in this study are available at the first author’s personal website: https://sites.google.com/view/tuan-d-pham/codes under the title “Prognostication of head injuries”.

## Funding

There was no funding for this work.

## Notes

### Competing Interest Statement

The authors have declared no competing interest.

### Funding Statement

This study did not receive any funding.

## References

[1] Dewan MC, Rattani A, Gupta S, Baticulon RE, Hung YC, Punchak M, Agrawal A, Adeleye AO, Shrime MG, Rubiano AM, Rosenfeld JV, Park KB. Estimating the global incidence of traumatic brain injury. J Neurosurg. 2018; 130(4):1080–1097. doi: 10.3171/2017.10.JNS17352.

[2] Maas AIR, Menon DK, Manley GT, Abrams M, Akerlund C, et al. Traumatic brain injury: progress and challenges in prevention, clinical care, and research. Lancet Neurol. 2022 Nov;21(11):1004–1060. doi: 10.1016/S1474-4422(22)00309-X.

[3] Brain health. WHO. https://www.who.int/health-topics/brain-health?utm\_source=chatgpt.com#tab=tab\_1. Accessed 15 January 2025.

[4] Traumatic Brain Injury & Concussion. CDC. https://www.cdc.gov/traumatic-brain-injury/data-research/index.html. Accessed 15 January 2025.

[5] GBD 2021 Nervous System Disorders Collaborators. Global, regional, and national burden of disorders affecting the nervous system, 1990-2021: a systematic analysis for the Global Burden of Disease Study 2021. Lancet Neurol. 2024; 23(4):344-381. doi: 10.1016/S1474-4422(24)00038-3.

[6] Falk H, Bechtold KT, Peters ME, Roy D, Rao V, Lavieri M, Sair H, Van Meter TE, Korley F. A prognostic model for predicting one-month outcomes among emergency department patients with mild traumatic brain injury and a presenting Glasgow Coma Scale of fifteen. J Neurotrauma. 2021; 38(19):2714–2722. doi: 10.1089/neu.2021.0137.

[7] Helmrich IRAR, van Klaveren D, Dijkland SA, Lingsma HF, Polinder S, Wilson L, von Steinbuechel N, van der Naalt J, Maas AIR, Steyerberg EW; CENTER-TBI Collaborators. Development of prognostic models for Health-Related Quality of Life following traumatic brain injury. Qual Life Res. 2022; 31(2):451–471. doi: 10.1007/s11136-021-02932-z.

[8] Patel S, Maria-Rios J, Parikh A, Okorie ON. Diagnosis and management of elevated intracranial pressure in the emergency department. Int J Emerg Med. 2023; 16(1):72. doi: 10.1186/s12245-023-00540-x.

[9] Jung MK, Roh TH, Kim H, Ha EJ, Yoon D, Park CM, Kim SH, You N, Kim GJ. Hyperosmolar therapy response in traumatic brain injury: Explainable artificial intelligence based long-term time series forecasting approach. Expert Systems with Applications 2024; 255 (part D): 124795, doi:10.1016/j.eswa.2024.124795.

[10] Sahuquillo J, Dennis JA. Decompressive craniectomy for the treatment of high intracranial pressure in closed traumatic brain injury. Cochrane Database Syst Rev. 2019; 12(12):CD003983. doi: 10.1002/14651858.CD003983.pub3.

[11] Grewal K, Atzema CL, Austin PC, de Wit K, Sharma S, Mittmann N, Borgundvaag B, McLeod SL. Intracranial hemorrhage after head injury among older patients on anticoagulation seen in the emergency department: a population-based cohort study. CMAJ 2021; 193(40):E1561–E1567. doi: 10.1503/cmaj.210811.

[12] Minhas H, Welsher A, Turcotte M, Eventov M, Mason S, Nishijima DK, Versmee G, Li M, de Wit K. Incidence of intracranial bleeding in anticoagulated patients with minor head injury: a systematic review and meta-analysis of prospective studies. Br J Haematol. 2018; 183(1):119–126. doi: 10.1111/bjh.15509.

[13] Furtan S, Pochcia-l P, Timler D, Ricci F, Sutton R, Fedorowski A, Zýsko D. Prognosis of syncope with head injury: a tertiary center perspective. Front Cardiovasc Med. 2020;7:125. doi: 10.3389/fcvm.2020.00125.

[14] Bressan S, Eapen N, Phillips N, Gilhotra Y, Kochar A, Dalton S, Cheek JA, Furyk J, Neutze J, Williams A, Hearps S, Donath S, Oakley E, Singh S, Dalziel SR, Borland ML, Babl FE; Paediatric Research in Emergency Departments International Collaborative (PREDICT). PECARN algorithms for minor head trauma: risk stratification estimates from a prospective PREDICT cohort study. Acad Emerg Med. 2021; 28(10):1124–1133. doi: 10.1111/acem.14308.

[15] Gizli G, Durak VA, Koksal O. The comparison of PECARN, CATCH, and CHALICE criteria in children under the age of 18 years with minor head trauma in emergency department. Hong Kong Journal of Emergency Medicine 2022; 29: 31-37. doi:10.1177/1024907920930510.

[16] Atis GM, Alta T, Atis SE. Comparison of CATCH, PECARN, and CHALICE clinical decision rules in pediatric patients with mild head trauma. Eur J Trauma Emerg Surg. 2022; 48(4):3123–3130. doi: 10.1007/s00068-021-01859-x.

[17] Ak R, Ç elik NB, Erdoǧan HD, Karakucuk AY, Gokdogan S, Korkmaz S, Seyhan AU. Evaluation of three clinical decision rules in pediatric patients with minor head injury: PECARN, CHALICE and CHATCH. Glob Emerg Crit Care 2023; 2(2):33–40. doi:10.4274/globecc.galenos.2023.69885.

[18] Bressan S, Kochar A, Oakley E, Borland M, Phillips N, Dalton S, Lyttle MD, Hearps S, Cheek JA, Furyk J, Neutze J, Dalziel S, Babl FE; Paediatric Research in Emergency Department International Collaborative (PREDICT) group. Traumatic brain injury in young children with isolated scalp haematoma. Arch Dis Child. 2019;104(7):664–669. doi: 10.1136/archdischild-2018-316066.

[19] Wan-Ting C, Chin-Hsien L, Cheng-Yu L, Cheng-Yu C, Chi-Chun L, Keng-Wei C, Jiann-Hwa C, Wei-Lung C, Chien-Cheng H, Cherng-Jyr L, Jui-Yuan C. Reverse shock index multiplied by Glasgow Coma Scale (rSIG) predicts mortality in severe trauma patients with head injury. Sci Rep. 2020; 10(1):2095. doi: 10.1038/s41598-020-59044-w.

[20] Ramdheen S, Naicker B. Evaluating the burden of head injuries on a rural emergency department in South Africa. S Afr Fam Pract (2004) 2021; 63(1):e1-e6. doi: 10.4102/safp.v63i1.5327.

[21] Aramaki E, Wakamiya S, Yada S, Nakamura Y. Natural language processing: from bedside to everywhere. Yearb Med Inform. 2022; 31(1):243–253. doi: 10.1055/s-0042-1742510.

[22] Alqahtani T, Badreldin HA, Alrashed M, Alshaya AI, Alghamdi SS, Bin Saleh K, Alowais SA, Alshaya OA, Rahman I, Al Yami MS, Albekairy AM. The emergent role of artificial intelligence, natural learning processing, and large language models in higher education and research. Res Social Adm Pharm. 2023; 19(8):1236–1242. doi: 10.1016/j.sapharm.2023.05.016.

[23] Oniani D, Chandrasekar P, Sivarajkumar S, Wang Y. Few-Shot Learning for Clinical Natural Language Processing Using Siamese Neural Networks: Algorithm Development and Validation Study. JMIR AI. 2023 May 4;2:e44293. doi: 10.2196/44293.

[24] Park HA, Jeon I, Shin S-H, Seo SY, Lee JJ, Kim C, Park JO. Natural language processing-based deep learning to predict the loss of consciousness event using emergency department text records. Applied Sciences 2024; 14(23):11399. doi:10.3390/app142311399.

[25] Yeung JA, Shek A, Searle T, Kraljevic Z, Dinu V, Ratas M, Al-Agil M, Foy A, Rafferty B, Oliynyk V, Teo JT. Natural language processing data services for healthcare providers. BMC Med Inform Decis Mak. 2024; 24(1):356. doi: 10.1186/s12911-024-02713-x.

[26] Pham TD. Classification of pediatric dental diseases from panoramic radiographs using natural language transformer and deep learning models. medRxiv 2025. doi:10.1101/2025.01.30.25321418.

[27] Khaki D, Hietanen V, Corell A, Herg’es HO, Ljungqvist J. Selection of CT variables and prognostic models for outcome prediction in patients with traumatic brain injury. Scand J Trauma Resusc Emerg Med. 2021; 29(1):94. doi: 10.1186/s13049-021-00901-6.

[28] Rocha TAH, Elahi C, Cristina da Silva N, Sakita FM, Fuller A, Mmbaga BT, Green EP, Haglund MM, Staton CA, Nickenig Vissoci JR. A traumatic brain injury prognostic model to support in-hospital triage in a low-income country: a machine learning-based approach. J Neurosurg. 2019; 132(6):1961–1969. doi: 10.3171/2019.2.JNS182098.

[29] Steyerberg EW, Mushkudiani N, Perel P, Butcher I, Lu J, McHugh GS, Murray GD, Marmarou A, Roberts I, Habbema JD, Maas AI. Predicting outcome after traumatic brain injury: development and international validation of prognostic scores based on admission characteristics. PLoS Med. 2008; 5(8):e165; discussion e165. doi: 10.1371/journal.pmed.0050165.

[30] Perel P, Edwards P, Wentz R, Roberts I. Systematic review of prognostic models in traumatic brain injury. BMC Med Inform Decis Mak. 2006; 6:38. doi: 10.1186/1472-6947-6-38.

[31] Hyder AA, Wunderlich CA, Puvanachandra P, Gururaj G, Kobusingye OC. The impact of traumatic brain injuries: a global perspective. NeuroRehabilitation 2007; 22(5):341–353.

[32] Lesko MM, Jenks T, Perel P, O’Brien S, Childs C, Bouamra O, Lecky F. Models of mortality probability in severe traumatic brain injury: results of the modelling by the UK trauma registry. J Neurotrauma. 2013; 30(24):2021–30. doi: 10.1089/neu.2013.2988.

[33] Hochreiter S, Schmidhuber J. Long short-term memory. Neural Comput. 1997; 9:1735-1780. doi:10.1162/neco.1997.9.8.1735.

[34] Devlin J, Chang MW, Lee K, Toutanova K. BERT: pre-training of deep bidirectional transformers for language understanding. In Proceedings of the 2019 Conference of the North American Chapter of the Association for Computational Linguistics: Human Language Technologies; 1:4171-4186 (2019).

[35] LeCun Y, Bottou L, Bengio Y, Haffner P. Gradient-based learning applied to document recognition. Proceedings of the IEEE 1998; 86(11):2278–2324. doi:10.1109/5.726791.

[36] Pham TD. Time-frequency time-space LSTM for robust classification of physiological signals. Sci Rep. 2021; 11(1):6936. doi: 10.1038/s41598-021-86432-7.

[37] Tu Z, Jeffries SD, Morse J, Hemmerling TM. Comparison of time-series models for predicting physiological metrics under sedation. J Clin Monit Comput. 2024. doi: 10.1007/s10877-024-01237-z.

[38] Gers FA, Schmidhuber E. LSTM recurrent networks learn simple context-free and context-sensitive languages,” in IEEE Transactions on Neural Networks 2001; 12(6): 1333–1340. doi: 10.1109/72.963769.

[39] Schmidhuber J, Gers F, Eck D. Learning nonregular languages: a comparison of simple recurrent networks and LSTM. Neural Comput. 2002; 14(9):2039–2041. doi: 10.1162/089976602320263980.

[40] Greff K, Srivastava RK, Koutńık J, Steunebrink BR. Schmidhuber J. LSTM: a Search space odyssey. IEEE Transactions on Neural Networks and Learning Systems 2017; 28(10): 2222–2232. doi: 10.1109/TNNLS.2016.2582924.

[41] Pham TD, Holmes SB, Zou L, Patel M, Coulthard P. Diagnosis of pathological speech with streamlined features for long short-term memory learning. Comput Biol Med. 2024; 170:107976. doi: 10.1016/j.compbiomed.2024.107976.

[42] Jamshidi S, Mohammadi M, Bagheri S, Najafabadi HE, Rezvanian A, Gheisari M, Ghaderzadeh M, Shahabi AS, Wu Z. Effective text classification using BERT, MTM LSTM, and DT. Data Knowl. Eng. 2024; 151:102306. doi: 10.1016/j.datak.2024.102306.

[43] Sun C, Yang Z, Wang L, Zhang Y, Lin H, Wang J. Biomedical named entity recognition using BERT in the machine reading comprehension framework. J Biomed Inform. 2021; 118:103799. doi: 10.1016/j.jbi.2021.103799.

[44] Duan K, Du S, Zhang Y, Lin Y, Wu H, Zhang Q. Enhancement of question answering system accuracy via transfer learning and BERT. Applied Sciences 2022; 12(22):11522. doi:10.3390/app122211522.

[45] Richardson K, Hu H, Moss L, Sabharwal A. Probing natural language inference models through semantic fragments. Proceedings of the AAAI Conference on Artificial Intelligence 2020; 34:8713–8721. doi:10.1609/aaai.v34i05.6397.

[46] Eleftheriadis P, Perikos I, Hatzilygeroudis I. Evaluating deep learning techniques for natural language inference. Applied Sciences 2023; 13(4):2577. doi:10.3390/app13042577.

